# Study of Coronavirus Impact on Parisian Population from April to June using Twitter and Text Mining Approach

**DOI:** 10.1101/2020.08.15.20175810

**Authors:** Josimar E. Chire Saire, Jimy Frank Oblitas Cruz

## Abstract

The fast spreading of coronavirus name covid19, generated the actual pandemic forcing to change daily activities. Health Councils of each country promote health policies, close borders and start a partial or total lockdown. One of the first countries in Europe with high impact was Italy. Besides at the end of April, one country with a shared border was on the top of 10 countries with more total cases, then France started with its own battle to beat coronavirus. This paper studies the impact of coronavirus in the poopulation of Paris, France from April 23 to June 18, using Text Mining approach, processing data collected from Social Network and using trends related of searching. First finding is a decreasing pattern of publications/interest, and second is related to health crisis and economical impact generated by coronavirus.

## 1 Introduction

Officially declared as a global pandemic by the World Health Organization (WHO) on March 11, 2020, COVID-19 outbreak (Coronavirus 19 disease) has evolved at an unprecedented rate. The COVID-19 pandemic has resulted in over 20 million confirmed cases and over 700,000 deaths globally. It has also sparked fears of an impending economic crisis and recession [1].

Social distancing, self-isolation and travel restrictions have lead to a reduced workforce across all economic sectors and caused many jobs to be lost. Schools have closed down, and the need for commodities and manufactured products has decreased. In contrast, the need for medical supplies has significantly increased.

All countries that have been affected by COVID 19 have followed a similar pandemic growth curve, where the number of cases of SARS-CoV-2 coronavirus infection continues to grow, and, as time goes by, together with prevention policies, the rate of contagion will start to decrease progressively until the situation is controlled. This has been observed in realities such as those of European countries, which shows that there is certain universality in the temporary evolution of COVID-19. This is demonstrated by the time lag graphs of infected populations confirmed in countries such as France, China and Italy, which follow the same power law on average [2].

In order to help public health and to make better decisions regarding Public Health and to help with their monitoring, Twitter has demonstrated to be an important information source related to health on the Internet, due to the volume of information shared by citizens and official sources. Twitter provides researchers an information source on public health, in real time and globally. Thus, it could be very important for public health research [3]

Within the context of COVID 19, users from all over the world may use it to identify quickly the main thoughts, attitudes, feelings and matters in their minds regarding this pandemic. This may help those in charge to make policies, health professionals and public in general to identify the main problems that concern everybody and deal with them more properly [4]

Particularly, focusing on a densely populated region of France, we document evidence that the highest economic “indicators of precariousness,” such as unemployment and poverty rates, lack of formal education and housing, are important factors in determining mortality rates for COVID-19. Therefore, measuring what happens after having the pandemic under control is essential, and the economic issue is important to be monitored, since it goes hand in hand with public health policies for the containment of the pandemic [5], so that our study will help to show changes in issues that concern the French population at this stage.

The actual paper uses Data Mining Approach to perform an exploratory analysis of the dataset of Brazilian patients of Sao Paulo State. The methodology to explore data is presented in Section 2, the experiments and results in Section 3. Conclusion states in Section 4, final recommendations and future work are presenten in Section 5, 6.

## 2 Methodology

The conducted work follows a methodology inspired in CRISP-DM[6]. This methodology is explained in the next subsections, from collecting data, processing and visualization to support the study.

### 2.1 Collecting data

Twitter is a Social Network, where users can post/share ideas, opinions, thoughts about any topic. Then, it is possible to collect text using Twitter API(Aplication Programming Interface). The parameters for accesing the data are:

– terms: covid19, coronavirus
– Date collection: 23/04/2020 – 18/06/2020
– Geolocalization: Paris, France
– Language: French
– Radius: 50 kilometers

### 2.2 Pre-processing Data

A cleaning process is necessary to avoid characters with no meaning for the scope of this analysis. First, convert text to lowercase, remove French accents, remove non alphanumerical values. Later, delete stopwords, i.e. articles, pronouns, etc.

### 2.3 Analysis

The scope of this paper is to analyze how reacted French population during the range of date: final week of May until third week of June. Besides, know how was the perception around Economy situation in Paris, France. This step is important to know what kind of graphics will be useful to answer the questions related to the study.

### 2.4 Visualization

The collected date is textual then filtering, organizing it to show proper graphics that support analysis and let a better understanding about the situation in Paris, France. Cloud of words are useful to get a general overview, bar plots for frequency or histograms, and filtering process removing some terms can help to get a better view of terms.

## 3 Experiments and Results

The present paper presents the description of dataset and results in the next subsections 3.1, 3.2. The results presents the interest of Parisian inhabitants about health crisis originated by coronavirus and concern around Econonomy topic.

### 3.1 Dataset

The collected data has the next features:

– Size: 1,496,375 tweets
– Fields: date(YYYY-MM-DD,HH:MM:SS), text(alphanumerical)
– Unique users: 285 114
– Date: 23/04/2020 – 18/06/2020

### 3.2 Results

All countries, including France, in response to ‘flattening the curve’, generated policies and rules for actions, such as border closures, travel restrictions and quarantine, which is a serious blow to one of Europe’s largest economies. These actions gave results, achieving a control of the epidemic, evidenced in the Figure 1, where it is clearly observed that since May a constant control of this one was achieved. [7].

**Fig. 1.**
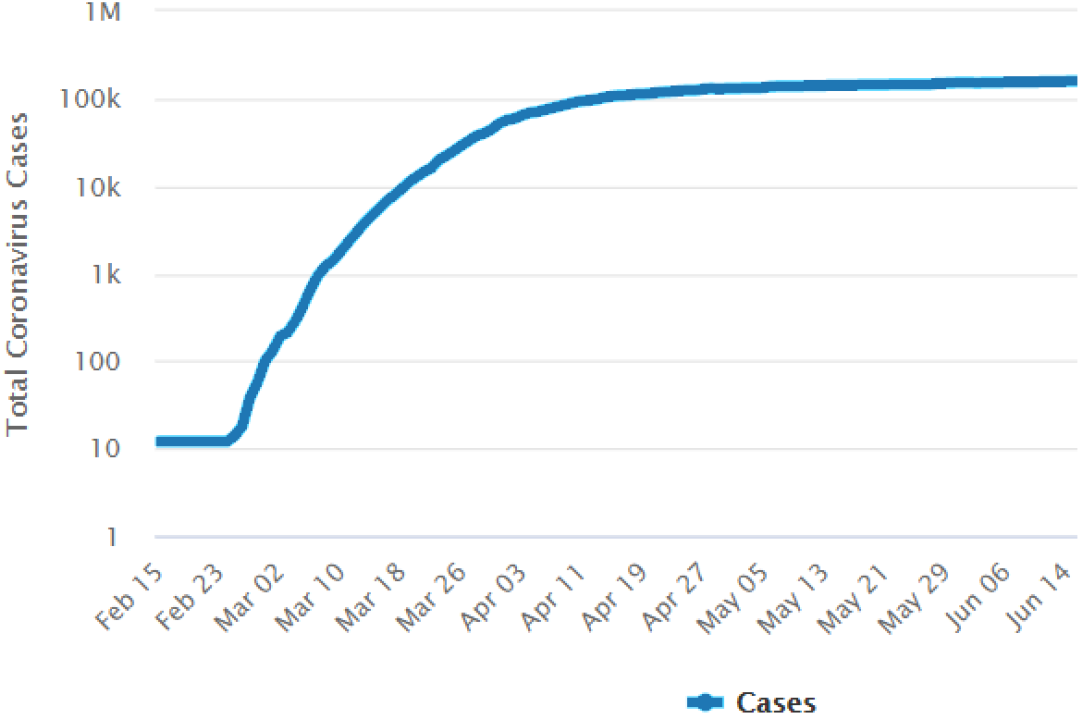
Total Cases France - Logarithmic Scale

A general ovierview of the collected is presented in Figure 2, after Fig. 3 presents the hourly distribution of downloaded Twitter posts. It is possible to appreciate that the process of downloading data recovered data from March to June 2020, where clearly the interest evidenced in the web begins to decrease with respect to topics related to fear of the disease, which was very high in previous periods. This assessment is based on the discussion about fear of COVID 19 on Twitter and the period in which the code to download the data was executed. The issues of fear of this pandemic have been related to issues of quarantine exhaustion, anxiety, depression and fear [8].

**Fig. 2.**
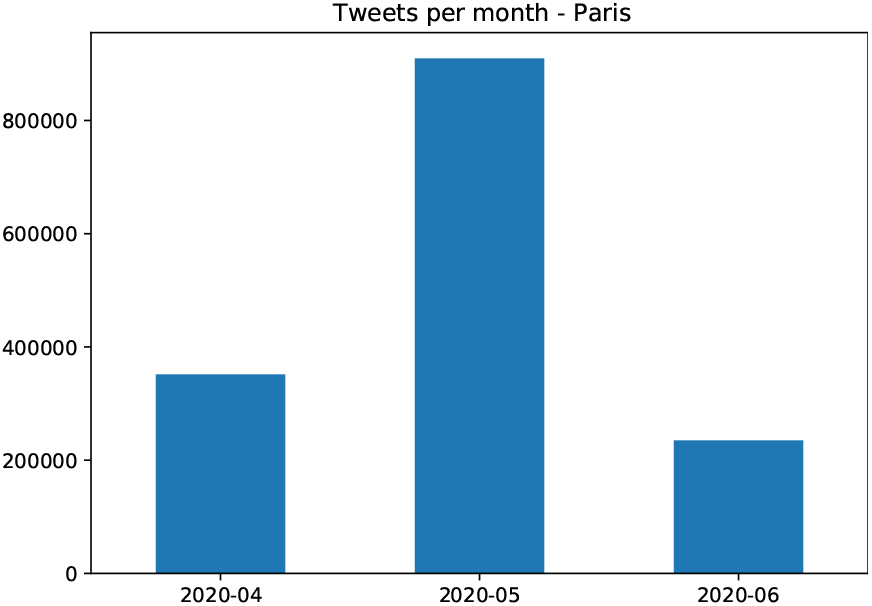
Number of tweets for month

**Fig. 3.**
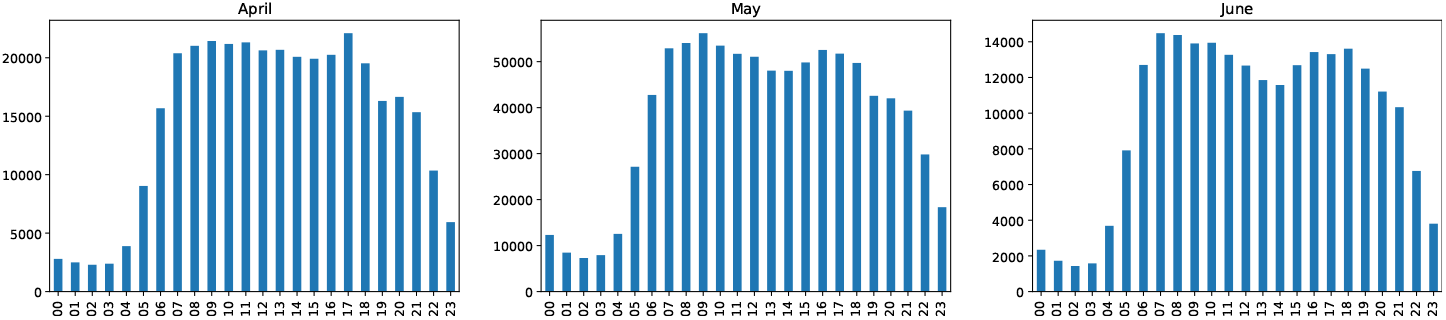
Number of tweets per hour, monthly

It is necessary to remark there is a clear pattern of publication in population of Paris from April to June(see 3). People start interaction at 6h, continues during noon passing afternoon and decreases from 19 – 21h. Besides, considering image 2, duplicating number of June to have an estimation of the total number for this month. There is a decreasing pattern of publications. By the other hand, a small valley is starting to appear around 13–14h, on May and June.

Enforcing the previous affirmation related to lose interest around covid19 topic, a combined graphic of Google Trends about coronavirus and number of publications with an adapted scale to have values between 0 to 100. An insight related to publications and interest is present in figure 4. Then people is losing interest on the pandemic, this can happen for many reasons.

**Fig. 4.**
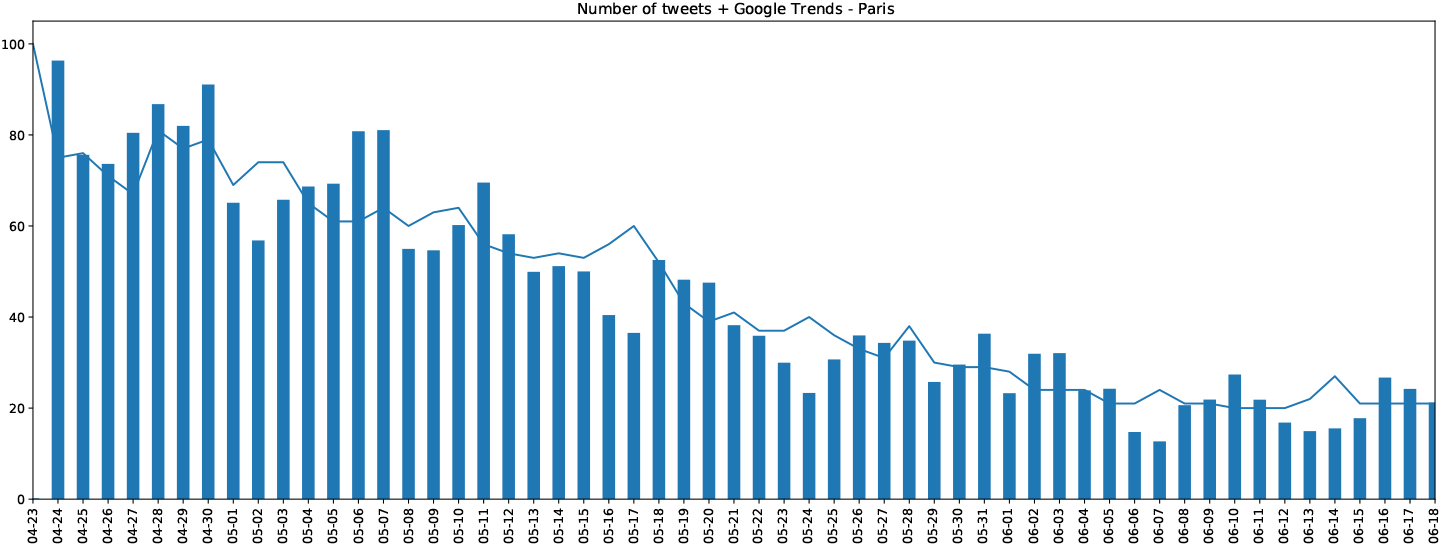
Number of tweets per day vs Google Trends

Helping the visualisation a cloud of words is presented in Fig. 5, and 6, it can be seen the regions including words related to “corp lutter”, “tue comment”, “plus parisien”, “plus personnes”, “Trump”, these tweets reflect the early interesting around the coronavirus health global crisis on this social network. All the collected data were searched using the keyword “Coronavirus”. Prior to the outbreak of COVID-19, people already relied on social media to gather information and news, and since the outbreak in January 2020, people in many countries have relied on social media like twitter to obtain information about the virus.

**Fig. 5.**
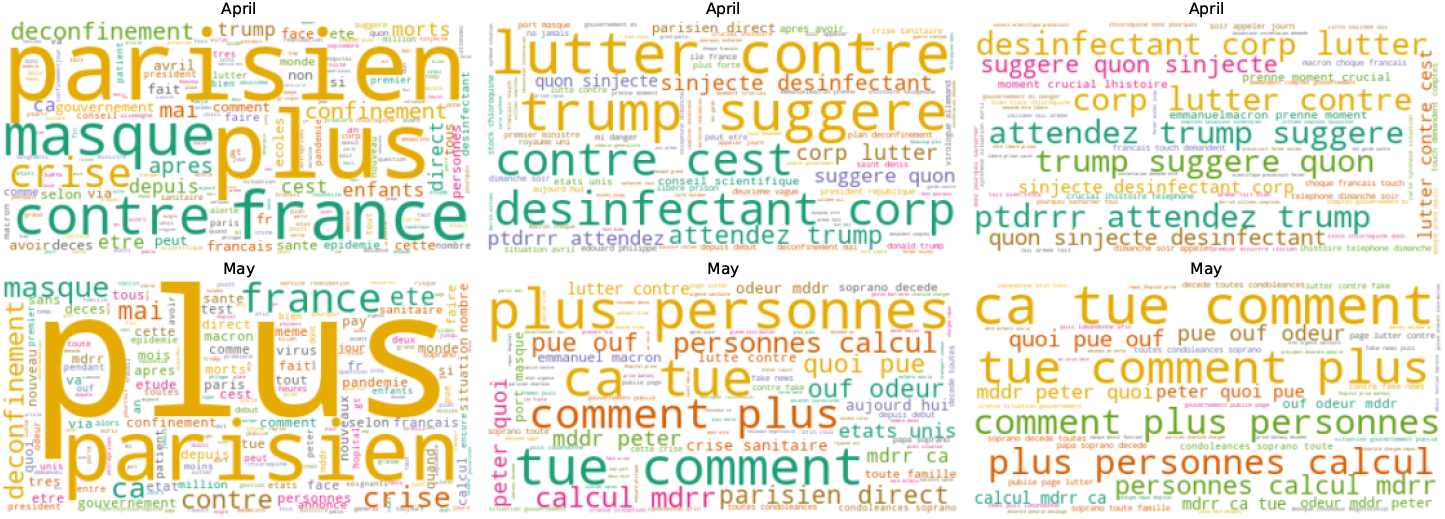
Cloud of words – April, May

**Fig. 6.**
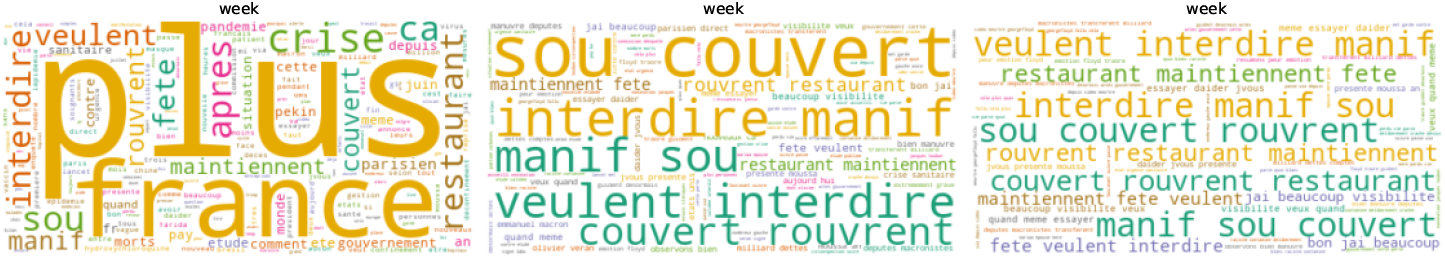
Cloud of words – June

But along with this, the emergence of new causes of anxiety, as detected in the words associated “crise”, “ouverture”, “pleine crise”, “crise économique” and “avant crise prix” (Fig. 7), with this analysis, is evident, being the main finding the fear of an imminent economic crisis and recession in France. the COVID-19 pandemic has had an unprecedented impact on the global economy as well as individuals’ economic well-being [9] [10] The shock of the coronavirus pandemic and shutdown measures to contain it have plunged the global economy into a severe contraction in countries where the pandemic has been the most severe and where there is heavy reliance on global trade, tourism, commodity exports, and external financing. According to World Bank forecasts, the global economy will shrink by 5.2 percent this year.[11]

**Fig. 7.**
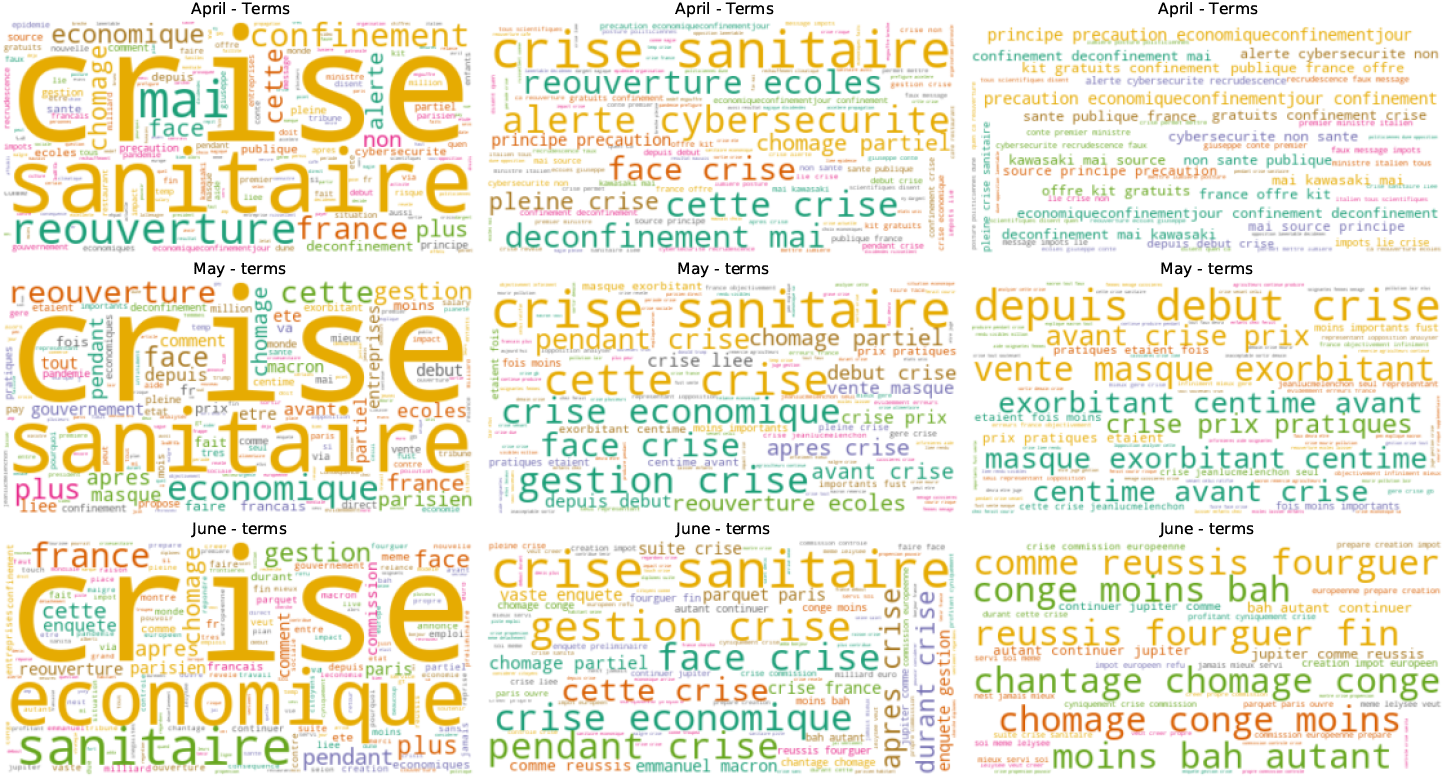
Cloud of words – April, May, June(Crisis)

This is reasonable as France’s leading newspapers already talked about the impact of COVID 19 on French economy and made efforts to try to understand the effect it would have, focusing on one of the most important sectors, which is tourism industry with all associated services [12], including impacts on both the supply and demand of travel[13]. As a direct consequence of COVID-19, the World Travel and Tourism Council warned that 50 million jobs in the global travel and tourism sector may be at risk [14].

The economic recessions are estimated to affect significantly on the people mental health and wellbeing by magnitude the relative and attributable risks. Research [15] indicates a significant adverse effect of job loss and unemployment on mental health sufferings like depression, stress, etc.

With this we can show that monitoring and using Text Mining techniques can detect changes in concerns and fears in the evolution of a population during and after the health emergency by COVID 19. This, along with the widespread popularity of social media that will provide the public with a fast platform to measure trends[16], makes this technique an important public health tool, as it measures in a short time the continuous evolution of communication strategies generated by government institutions.

## 4 Conclusions

The information analysis on Twitter indicated by the detected rates can help to monitor the evolution of the interests of a population like that of France, within the phase of control of the outbreak of the current COVID-19 pandemic, showing that public interest in fear of health issues decreased and new fears arose, such as the issue of economic crisis, which is relevant information to generate effective communication policies meeting the needs of a population within the framework of Public Health.

## Data Availability

Not applicable

## 5 Recommendations

For researchers interested to work with this approach, consider:

– Select a topic to study and check if Social Networks are a source for your work. Every country has different number of active users and preferences about Social Networks.
– Consider to use some tool to get an overview of geographical zone before of using a data collection of some city/state.
– Remember languages has patterns about how writing(grammar) besides slang or common phrases are dependant of the location, then if you can find one collaborator from the zone of study, this will be very valuable to support the analysis.
– Involve more people to avoid bias for your own thoughts/ideas and of course, invite specialists around the topic of analysis, they will give you the key terms, intuition about what is useful or not and enforce the project.

## 6 Future Work

Perform a deeper analysis about topics related to main sectors: Economy, Social, Health, Education. Invite collaborators, i.e. Economist, Sociologist, Physicists, Teachers to do a global analysis, how one sector can impact/influence to anthers as chain effect.

